# Respiratory virus detection and sequencing from negative SARS-CoV-2 rapid antigen tests

**DOI:** 10.1101/2024.08.19.24311993

**Authors:** Emmanuela Jules, Charlie Decker, Brianna Jeanne Bixler, Alaa Ahmed, Zijing (Carol) Zhou, Itika Arora, Henok Tafesse, Hannah Dakanay, Andrei Bombin, Ethan Wang, Jessica Ingersoll, Kathy Bifulco, Jennifer K. Frediani, Richard Parsons, Julie Sullivan, Morgan Greenleaf, Jesse J. Waggoner, Greg S. Martin, Wilbur A. Lam, Anne Piantadosi

**Author notes:** **Corresponding author:** Anne Piantadosi MD, PhD, Woodruff Memorial Research Building, 101 Woodruff Circle, Atlanta GA 30322. These first authors contributed equally to this article (Emmanuela Jules, Charlie Decker, Brianna Jeanne Bixler).

## Abstract

Genomic epidemiology offers important insight into the transmission and evolution of respiratory viruses. We used metagenomic sequencing from negative SARS-CoV-2 antigen tests to identify a wide range of respiratory viruses and generate full genome sequences, offering a streamlined mechanism for broad respiratory virus genomic surveillance.

## Introduction

The SARS-CoV-2 pandemic highlighted the importance of genomic epidemiology in understanding virus transmission and evolution, informing essential countermeasures from non-pharmaceutical interventions to vaccines. Massive global efforts in SARS-CoV-2 genomic surveillance were made possible by widespread diagnostic testing and the growth of new infrastructure and methods for sequencing and analysis (1). Most genomic surveillance pipelines in the U.S. obtained residual SARS-CoV-2 positive samples from clinical, public health, and commercial laboratories. This strategy was effective during the pandemic but difficult to maintain with the rise of at-home rapid antigen tests (2, 3). As traditional sample sources declined, our group and others demonstrated that residual samples from rapid antigen tests could be used to generate and analyze full SARS-CoV-2 sequences for genomic surveillance (4-6).

Here, we build upon this work by identifying, sequencing, and analyzing other respiratory viruses using residual swab samples from negative BinaxNow™ COVID-19 antigen tests. This multi-virus approach is important as SARS-CoV-2 has transitioned to an endemic virus whose symptoms resemble those of other respiratory viruses (7). Thus, there is both a need for broad testing and an opportunity to expand genomic surveillance for respiratory viruses using self-collected samples.

## Methods

Detailed laboratory and analysis methods are provided in the **Appendix**. Briefly, participants were enrolled in a parent study evaluating novel viral diagnostic tests through the RADx program at the Atlanta Center for Microsystems Engineered Point-of-Care Technologies. The study protocol was approved by the Emory Institutional Review Board and the Grady Research Oversight Committee. We performed RNA metagenomic sequencing as described (8), obtaining a median of 5.8 million reads per sample (**Supplementary Data**). We used a three-step bioinformatic approach to detect viruses (**Supplementary Figure 1**) using KrakenUniq, blastn, and reference mapping, with a final criterion requiring coverage of at least 3 distinct genome regions, based on clinical diagnostic criteria for metagenomic sequencing (9).

## Results

We collected negative BinaxNOW™ test samples from 53 individuals between April-August 2023 (**Supplementary Table 1)**, a period during which 68% of the BinaxNOW™ tests in the parent study were negative. All individuals were symptomatic at the time of testing (**Table 1)**, and the median interval between symptom onset and testing was 2 days (range 0-9). RT-PCR was positive for influenza B in three samples and negative for influenza A and SARS-CoV-2 in all samples (**Supplementary Data**).

**Table 1:** Participant symptoms. Table indicates the number and percent of participants reporting each symptom at the time of testing. For symptom categories (gray rows), the number of participants with at least one symptom in that category is reported. *This includes one individual with SARS-CoV-2 detected at a low level and 17 individuals with an alternative human pathogenic respiratory virus detected.

| Symptom | Total participants<br>(N = 53) | Participants with a<br>virus detected<br>(N = 18*) | Participants with no<br>virus detected<br>(N = 35) |
| --- | --- | --- | --- |
| **Upper Respiratory** | 47 (88.7%) | 17 (94.4%) | 30 (85.7%) |
| Congestion/runny nose | 37 (69.8%) | 15 (83.3%) | 22 (62.9%) |
| Sore throat | 33 (62.3%) | 14 (77.8%) | 19 (54.3%) |
| Loss of sense of taste or<br>smell | 7 (13.2%) | 2 (11.1%) | 5 (14.3%) |
| **Lower Respiratory** | 43 (81.1%) | 15 (83.3%) | 28 (80.0%) |
| Cough | 39 (73.6%) | 15 (83.3%) | 24 (68.6%) |
| Shortness of breath | 23 (43.4%) | 6 (33.3%) | 17 (48.6%) |
| **Gastrointestinal** | 15 (28.3%) | 6 (33.3%) | 9 (25.7%) |
| Vomiting | 4 (7.6%) | 3 (16.7%) | 1 (2.9%) |
| Nausea | 11 (20.8%) | 2 (11.1%) | 9 (25.7%) |
| Diarrhea | 2 (3.8%) | 1 (5.6%) | 1 (2.9%) |
| Abdominal pain | 7 (13.2%) | 2 (11.1%) | 5 (14.3%) |
| **Systemic** | 35 (66.0%) | 13 (72.2%) | 22 (62.9%) |
| Fever (>100.4) | 18 (34.0%) | 7 (38.9%) | 11 (31.4%) |
| Chills | 24 (45.3%) | 11 (61.1%) | 13 (37.1%) |
| Fatigue | 27 (50.9%) | 11 (61.1%) | 16 (45.71%) |
| **Other** | 41 (77.4%) | 16 (88.9%) | 25 (71.4%) |
| Headache | 30 (56.6%) | 13 (72.2%) | 17 (48.6%) |
| Joint pain | 14 (26.4%) | 3 (16.7%) | 11 (31.4%) |
| Muscle pain | 31 (58.5%) | 10 (55.6%) | 21 (60.0%) |

Metagenomic sequencing identified a low level of SARS-CoV-2 in one sample and a different pathogenic human respiratory virus in 17 of the other 52 samples (33%) (**Supplementary Data)**. The following viruses were detected: parainfluenza viruses (N=7), rhinoviruses (N=5), influenza B (N=3), seasonal coronaviruses (N=2), and adenovirus (N=1) (**Figure 1**). In one sample, both influenza B and parainfluenza 2 were detected. In another sample positive for influenza B by RT-PCR, metagenomic sequencing did not identify influenza but identified human mastadenovirus E. Thus, excluding SARS-CoV-2, a total of 18 viruses were detected across 17 samples. There was no difference in the total number of reads obtained for samples with and without viruses detected (Mann Whitney U test, p=0.29).

**Figure 1:**
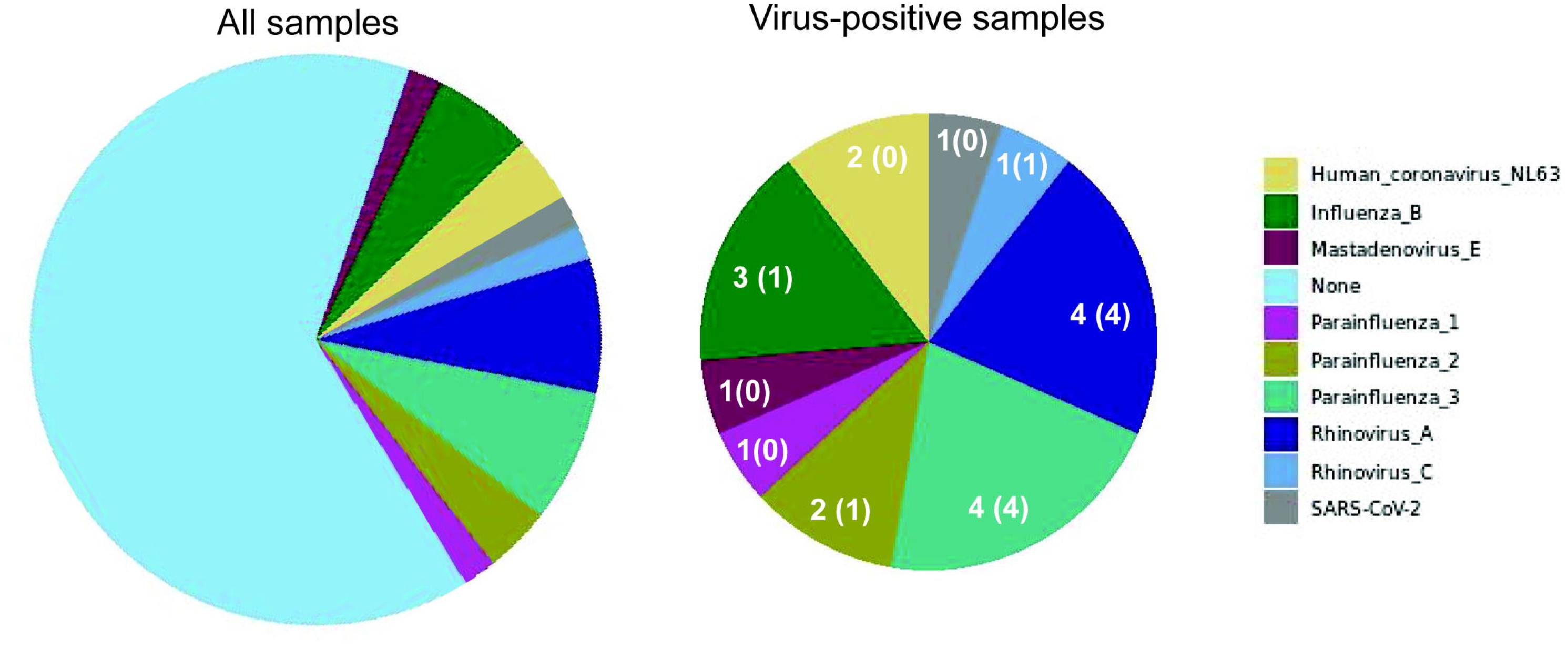
Frequency of human pathogenic respiratory viruses found in 53 residual samples from BinaxNOW™ tests that were negative for SARS-CoV-2. Pie charts indicate the number of samples positive for each virus among all samples (left panel) and among the 18 positive samples (right panel). Numbers indicate the number of samples with a virus identified, followed in parentheses by the number of samples with a >90% complete genome sequence assembled.

We observed potential differences in symptom frequencies between individuals with and without viruses detected, but none were statistically significant (**Table 1**).

Of the 18 viruses detected, we generated full viral genome sequences from 11 (61%), with >90% coverage and 71-24,000 fold depth (**Supplementary Data**). These included parainfluenza 3 (4/4 samples), parainfluenza 2 (1/2), rhinovirus (5/5), and influenza B (1/3).

We performed phylogenetic analysis of parainfluenza 3 as a proof-of-concept for genomic epidemiology studies and found substantial diversity. Using the lineage classification system described in (10), two of our sequences clustered with Lineage A1 sequences from 2019-2023 (**Figure 2A**), another clustered with Lineage C sequences from Japan in 2023, and the fourth with Lineage C sequences from the U.S. collected between 2015-2017 (**Figure 2B**), all with high bootstrap support (**Supplementary Figure 2**). Of note, there are only about 450 complete parainfluenza 3 virus sequences available; the data from our small study represent nearly 1% of this number, underscoring the opportunity to easily expand genomic surveillance using this approach.

**Figure 2:**
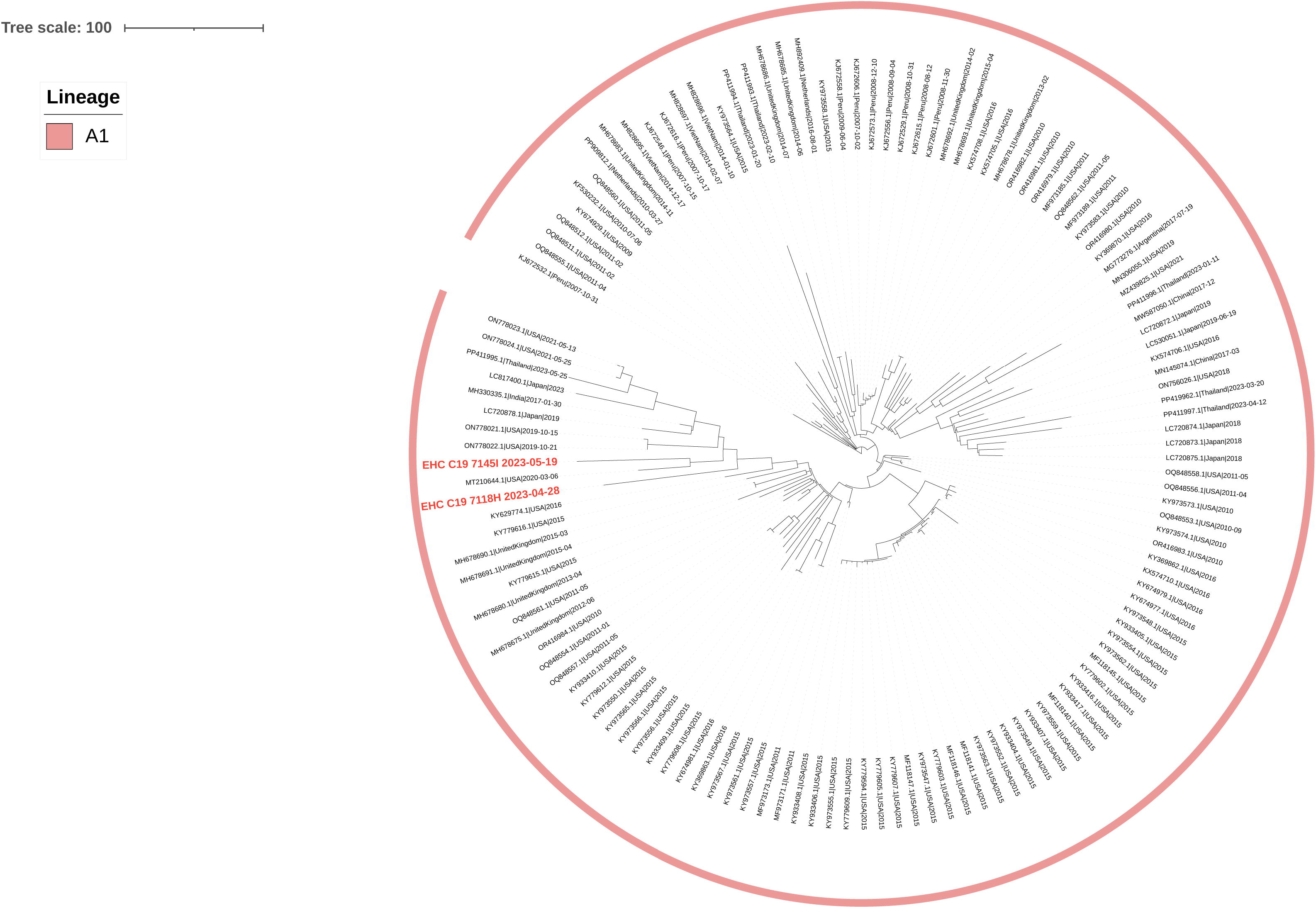
Phylogenetic analysis of parainfluenza 3 virus sequences. The names of sequences obtained in this study are bold and in red, and reference sequences are in black. The outer ring indicates virus lineage: **A)** contains representative lineage A1 sequences, and **B)** contains representative sequences from lineages C, E, F, and G. Each tree is a maximized parsimony subtree using down sampled data from the full analysis in **Supplementary Figure 2**, for ease of visualization.

In addition to human pathogenic respiratory viruses, we detected over 100 viruses of no clinical significance, including bacteriophages and plant viruses, many of which were also detected in our negative controls (**Figure 3**). Similarly, mastadenovirus C was found in many samples and negative controls. These are all consistent with environmental or reagent contaminants. Herpesviruses were found in many samples by KrakenUniq and blastn, but were not confirmed by mapping to a reference sequence with coverage of at least 3 regions. Overall, 1,367 viral taxa were identified by KrakenUniq, only 254 (18.6%) were confirmed by BLAST, and only 137 (53.9% of these, 10% of total) met our criteria for detection, highlighting the importance of confirmatory steps in metagenomic analysis.

**Figure 3:**
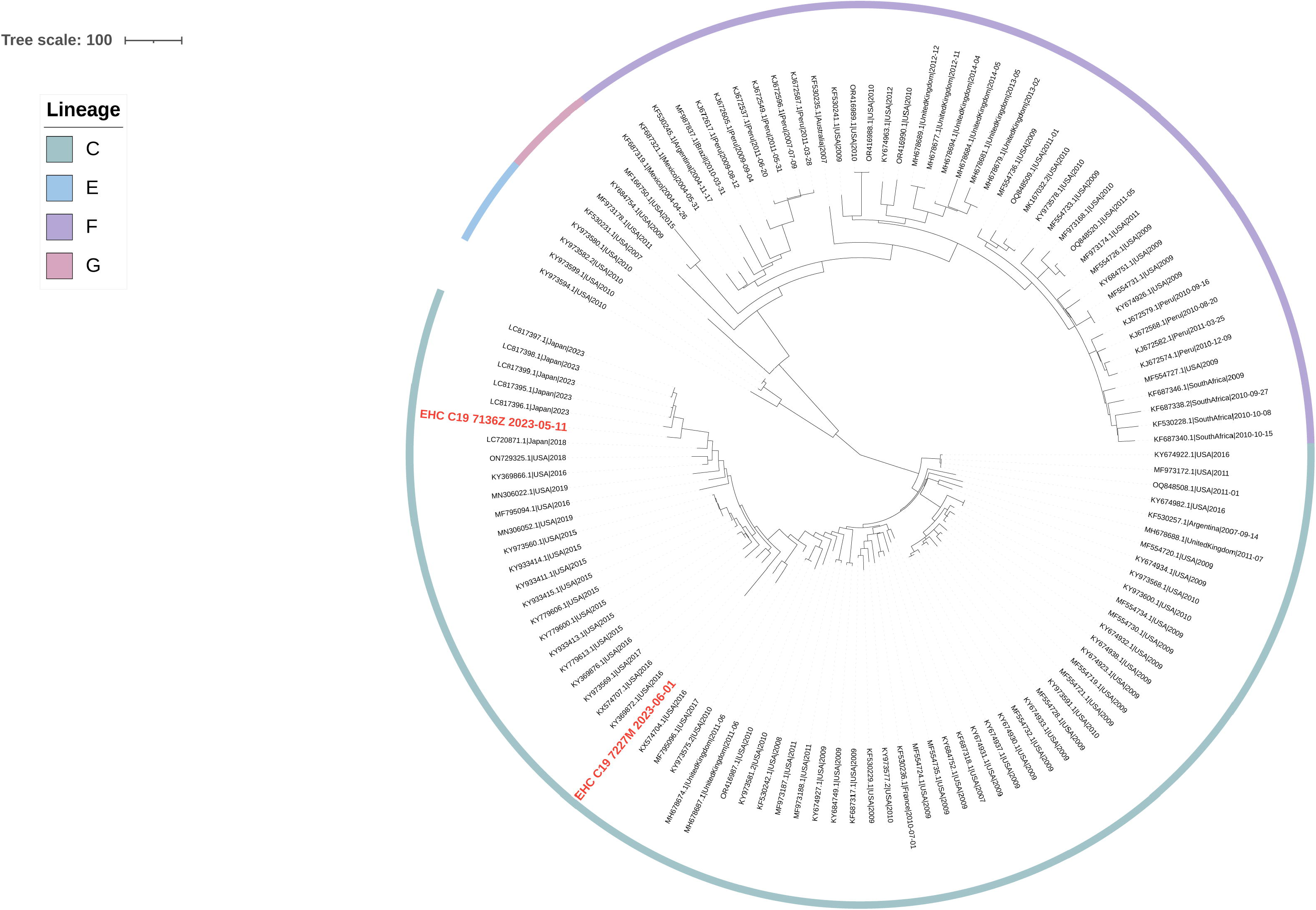
Plot of the viral taxa (rows) that were detected in each sample (columns). Blue boxes indicate viruses that were detected by both KrakenUniq and blastn but not confirmed by reference mapping, while red boxes indicate viruses that were detected by both KrakenUniq and blastn and were confirmed by reference mapping (covering at least 3 distinct regions of the reference virus genome).

## Discussion

Our study demonstrates that RNA metagenomic sequencing of residual swab samples from negative BinaxNOW™ tests can be used to detect a broad range of respiratory viruses, including rhinoviruses, parainfluenza viruses, influenza B, seasonal coronaviruses, and adenovirus. All of these have overlapping symptoms with one another and with SARS-CoV-2, underscoring the need for multi-virus testing approaches. Although our study was not designed for clinical diagnosis, metagenomic sequencing is increasingly used clinically, and our results illustrate the need for rigorous analysis techniques and careful interpretation.

It is notable that only 33% of samples had a human pathogenic respiratory virus. This is similar to our prior study detecting alternative respiratory viruses in only 40% of SARS-CoV-2 negative individuals using residual clinical samples early in the pandemic (8). Possible explanations include individuals with a non-infectious syndrome, a bacterial or other non-viral infection, or a virus present at a low level. It is also possible that some individuals were infected with a DNA virus not optimally captured by RNA sequencing. However, we detected adenovirus, the most prevalent respiratory DNA virus. Among common RNA viruses, we did not detect influenza A or RSV, which we attribute to the winter-predominant seasonality of these viruses compared to our sample collection in spring and summer.

Importantly, of the 18 viruses detected, we were able to generate full viral genome sequences from 11 (61%) using moderate sequencing depths. Thus, the single laboratory technique of metagenomic sequencing can not only identify diverse respiratory viruses but also contribute to their genomic surveillance. The surprisingly high depth of genome coverage achieved for many sequences indicates that throughput and cost can be improved by reducing total sequencing reads from each sample in future studies.

By combining metagenomic sequencing with the use of residual antigen test samples, we demonstrate a mechanism for convenient and broad respiratory virus surveillance. Our study used BinaxNOW™ tests, which conveniently preserve the used swab within the kit cassette; future work is needed to evaluate this approach using rapid antigen test strips themselves, as previously demonstrated for SARS-CoV-2 sequencing (5). Additionally, future studies would benefit from a regulatory framework in which results can be returned to study participants, who are likely curious about the presence of other respiratory viruses when rapid antigen testing is negative.

In conclusion, our study illustrates that residual samples from self-collected antigen tests can be a powerful sample source for investigating the genomic epidemiology of a broad range of respiratory viruses, building upon the strong foundations for viral surveillance established during the SARS-CoV-2 pandemic.

## Data Availability

All raw sequencing data (cleaned of human reads) is available in NCBI SRA under BioProject PRJNA634356, and assembled virus genome sequences are available in NCBI GenBank with accession numbers listed in the Supplementary Data file.

## Data Availability

All raw sequencing data (cleaned of human reads) is available in NCBI SRA under BioProject PRJNA1144955, and assembled virus genome sequences are available in NCBI GenBank with accession numbers listed in the Supplementary Data file.

## Acknowledgements

We would like to thank the study participants.

## Author bio

Ms. Jules received a Bachelor of Science in Anthropology and Human Biology from Emory University and is currently a research specialist in the Department of Pathology and Laboratory Medicine in the Emory University School of Medicine. She will be applying to medical school with the aspiration of becoming a family doctor and expanding healthcare to underserved communities.

## Figures

**Supplementary Figure 1:**
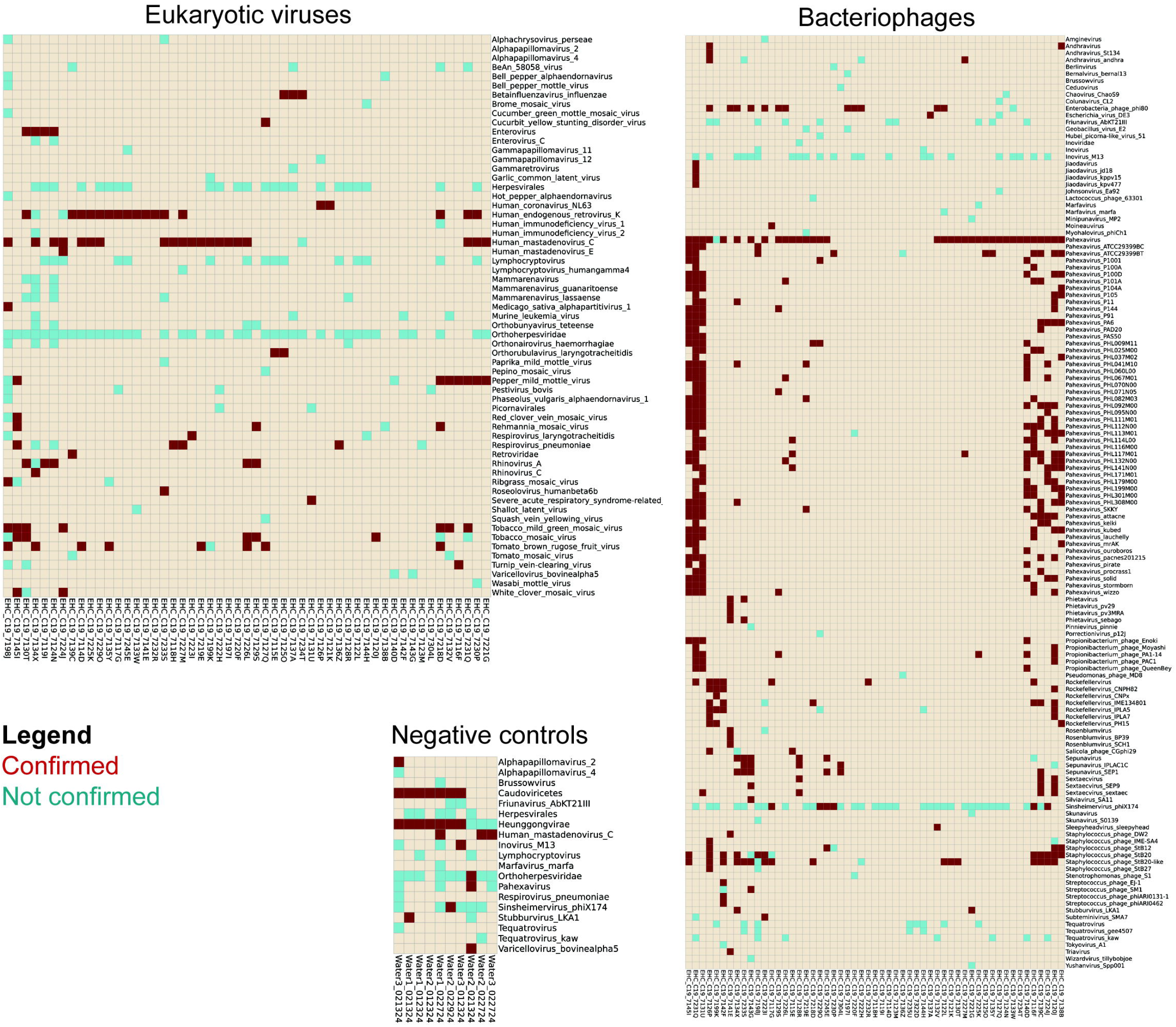
Analysis pipeline for metagenomic classification and confirmation. Red boxes describe processing and quality control steps, yellow boxes describe initial metagenomic classification using KrakenUniq, green boxes describe blastn confirmation of reads classified as viral by KrakenUniq, and blue boxes describe reference-based mapping for final confirmation. LCA = lowest common ancestor.

**Supplementary Figure 2: Maximum likelihood phylogenetic analysis of parainfluenza 3 virus sequences**. The names of sequences obtained in this study are bold and in red, and reference sequences in black represent all unique full-length genome sequences of parainfluenza 3 available in GenBank (7/30/24). Circles indicate nodes with >95% ultrafast bootstrap support. The outer ring indicates virus lineage.

## References

1. Oude Munnink BB, Worp N, Nieuwenhuijse DF, Sikkema RS, Haagmans B, Fouchier RAM, et al. The next phase of SARS-CoV-2 surveillance: real-time molecular epidemiology. Nat Med. 2021;27(9):1518–24. Epub 20210909. doi: 10.1038/s41591-021-01472-w. PubMed PMID: 34504335.

2. Ritchey MD, Rosenblum HG, Del Guercio K, Humbard M, Santos S, Hall J, et al. COVID-19 Self-Test Data: Challenges and Opportunities - United States, October 31, 2021-June 11, 2022. MMWR Morb Mortal Wkly Rep. 2022;71(32):1005–10. Epub 20220812. doi: 10.15585/mmwr.mm7132a1. PubMed PMID: 35951486; PubMed Central PMCID: PMC9400539.

3. Rader B, Gertz A, Iuliano AD, Gilmer M, Wronski L, Astley CM, et al. Use of At-Home COVID-19 Tests - United States, August 23, 2021-March 12, 2022. MMWR Morb Mortal Wkly Rep. 2022;71(13):489–94. Epub 20220401. doi: 10.15585/mmwr.mm7113e1. PubMed PMID: 35358168; PubMed Central PMCID: PMC8979595.

4. Nguyen PV, Carmola LR, Wang E, Bassit L, Rao A, Greenleaf M, et al. SARS-CoV-2 molecular testing and whole genome sequencing following RNA recovery from used BinaxNOW COVID-19 antigen self tests. J Clin Virol. 2023;162:105426. Epub 20230324. doi: 10.1016/j.jcv.2023.105426. PubMed PMID: 37028004; PubMed Central PMCID: PMC10036152.

5. Martin GE, Taiaroa G, Taouk ML, Savic I, O’Keefe J, Quach R, et al. Maintaining genomic surveillance using whole-genome sequencing of SARS-CoV-2 from rapid antigen test devices. Lancet Infect Dis. 2022;22(10):1417–8. Epub 20220804. doi: 10.1016/S1473-3099(22)00512-6. PubMed PMID: 35934015; PubMed Central PMCID: PMC9352270.

6. Hassouneh SA, Trujillo A, Ali S, Cella E, Johnston C, DeRuff KC, et al. Antigen test swabs are comparable to nasopharyngeal swabs for sequencing of SARS-CoV-2. Sci Rep. 2023;13(1):11255. Epub 20230712. doi: 10.1038/s41598-023-37893-5. PubMed PMID: 37438412; PubMed Central PMCID: PMC10338537.

7. Geismar C, Nguyen V, Fragaszy E, Shrotri M, Navaratnam AMD, Beale S, et al. Symptom profiles of community cases infected by influenza, RSV, rhinovirus, seasonal coronavirus, and SARS-CoV-2 variants of concern. Sci Rep. 2023;13(1):12511. Epub 20230802. doi: 10.1038/s41598-023-38869-1. PubMed PMID: 37532756; PubMed Central PMCID: PMC10397315.

8. Babiker A, Bradley HL, Stittleburg VD, Ingersoll JM, Key A, Kraft CS, et al. Metagenomic Sequencing To Detect Respiratory Viruses in Persons under Investigation for COVID-19. J Clin Microbiol. 2020;59(1). Epub 20201217. doi: 10.1128/JCM.02142-20. PubMed PMID: 33067271; PubMed Central PMCID: PMC7771462.

9. Miller S, Naccache SN, Samayoa E, Messacar K, Arevalo S, Federman S, et al. Laboratory validation of a clinical metagenomic sequencing assay for pathogen detection in cerebrospinal fluid. Genome Res. 2019;29(5):831–42. Epub 20190416. doi: 10.1101/gr.238170.118. PubMed PMID: 30992304; PubMed Central PMCID: PMC6499319.

10. Lee K, Park K, Sung H, Kim MN. Phylogenetic lineage dynamics of global parainfluenza virus type 3 post-COVID-19 pandemic. mSphere. 2024;9(4):e0062423. Epub 20240319. doi: 10.1128/msphere.00624-23. PubMed PMID: 38501829; PubMed Central PMCID: PMC11036794.

