## Appendix for "Respiratory virus detection and sequencing from negative SARS-CoV-2 rapid antigen tests"

### Supplementary Methods

#### Participants and clinical data

Participants were enrolled in a parent study to evaluate novel viral diagnostic tests as part of the RADx program at the Atlanta Center for Microsystems Engineered Point-of-Care Technologies (ACME-POCT). The study protocol was approved by the Emory Institutional Review Board and the Grady Research Oversight Committee. Participants were enrolled if symptomatic, and they performed testing in a monitored setting to ensure the test was performed correctly. We collected residual negative BinaxNOW™ test samples from 53 individuals between April-August 2023. Used cassettes containing swabs were stored at 4°C in sealed bags for a median of 6 days (range 2 - 19). Demographic and clinical data were collected in a secure database (REDCap, Nashville, TN). Symptom frequencies were compared between groups (individuals with a virus detected vs. those without) using chi-squared tests with Yates correction, or Fisher's exact test for comparisons with small numbers.

#### Metagenomic RNA sequencing

Nucleic acid extraction was performed using the Qiagen EZ1 DSP Virus Kit with the Advanced XL instrument, per the manufacturer's instructions. Multiplex RT-PCR was performed for Influenza A and B and SARS-CoV-2 using a published assay(7). Metagenomic RNA sequencing was performed by DNase treatment (ArcticZymes), random primer first-strand cDNA synthesis using SuperScript IV (Invitrogen), and second-strand synthesis using RNase H, DNA polymerase, and ligase (NEB). Libraries were prepared using the Nextera XT kit (Illumina) and sequenced using an Illumina MiSeq or NextSeq 2000. A median of 5.8 million reads per sample was obtained (range 0.6-40.2 million, **Supplementary Data**). Ten water samples were included as negative controls.

**Metagenomic analysis**

We used a three-step bioinformatic approach to detect and confirm viruses in each sample (**Supplementary Figure 1**). Reads first underwent deduplication using clumpify v39.06, quality trimming and adapter removal using Trimmomatic v0.39 (4-base wide sliding window, minimum average quality of 15, leading and trailing base removal quality below 3), and further quality filtering and merging of paired reads using Fastp v0.23.2 (default settings, minimum length 50). Filtered reads underwent metagenomic classification with KrakenUniq v0.5.7, and viral reads were confirmed with blastn v2.12.0 against the BLAST nucleotide database Version 4. A custom python script used the NCBI taxonomy utility (updated 4/22/24) to determine the lowest common ancestor (LCA) between the top three results from KrakenUniq and blastn; if no viral LCA was identified, the read was designated a false-positive result from KrakenUniq.

For viral taxa confirmed by both KrakenUniq and blastn, the closest matching reference sequence in GenBank was identified using a custom script, as follows. This script, requiring the NCBI Taxonomy ID (taxID), first downloads genomic data from NCBI using the datasets download genome taxon command, saving it as tax_$taxID.zip and extracting it into dir_$taxID. It then concatenates all FASTA files in the extracted directory into a single file, all_$taxID.fna, by generating a list of .fna files and appending their contents. Finally, the script uses the makeblastdb command to create a nucleotide BLAST database named candidate_virus from the consolidated FASTA file. This pipeline ensures efficient genomic data processing and BLAST database preparation for subsequent bioinformatics analyses.

The next script in the pipeline processes zipped paired-end reads (`R1` and `R2`), concatenates them into a single FASTQ file, and converts this into a FASTA file. It then filters the sequences based on a provided CSV file, extracting only the blast-verified reads. The resulting subset of sequences is saved in a new FASTA file for further analysis.

The final script in the pipeline processes input files to identify and analyze sequence data against a BLAST database. It begins by running a BLAST search on the input FASTA file (`ffn`) against the specified BLAST database (`blastDB`), outputting the results in tabular format. The script identifies the reference sequence with the highest number of positive blast hits above an 80% identity threshold and generates a BED file listing the aligned regions. It then subsets the original FASTQ file to include only reads aligning to this reference sequence. The script calculates the percent coverage of the reference sequence by comparing the lengths of aligned regions and the reference sequence itself. Finally, it generates an output file summarizing the read count, reference sequence, taxonomy ID, region count, aligned region length, reference length, and percent coverage. This automated pipeline facilitates the detailed analysis of sequence data, providing insights into genomic alignments and coverage.

Reference-based assembly was performed using viralrecon v2.6.0. A virus was confirmed as present in a sample if there were reads covering at least 3 distinct regions of the genome, a threshold based on clinical diagnostic studies using metagenomic sequencing. Viruses that were found in multiple study samples and negative controls were regarded as environmental or reagent contaminants, including mastadenovirus C, which was found at low levels (<5% genome coverage) in 20 samples (38%) and 3 controls (30%).

**Phylogenetic analysis**

Phylogenetic analysis was performed for parainfluenza 3: reference sequences were obtained from GenBank (N=451) using the name “human respirovirus 3” (taxid 11216) and following conditions: sequence length 14,000-16,000 nucleotides, complete genomes, and human hosts (taxid 9606). Identical sequences were removed, leading to a final alignment of 423 reference sequences. The four parainfluenza 3 sequences obtained in this study were aligned with the 423 references and NC_075446.1 using MAFFT v7.505. A maximum likelihood phylogenetic tree was constructed with IQ-TREE v2.3.5, with the GTR+F+I+R5 model and 1000 ultrafast bootstrap replicates. For visualization purposes, the tree was downsampled using UShER to generate subtrees with reference sequences closely related to the sequences from this study. All trees were visualized and annotated with iTOL v6.9.1.

### Supplementary Table 1: Participant demographics

| | Total Participants = 53 |
|---|---|
| **Sex** | |
| Male | 17 (32.1%) |
| Female | 36 (66.1%) |
| **Race** | |
| Black or African American | 37 (69.8%) |
| White | 7 (13.2%) |
| Asian | 2 (3.7%) |
| Native Hawaiian or Other Pacific Islander | 0 (0.0%) |
| American Indian / Alaskan Native | 0 (0.0%) |
| Other | 6 (11.3%) |
| Unknown | 1 (1.9%) |
| **Ethnicity** | |
| Hispanic or Latino | 4 (7.6%) |
| Not Hispanic or Latino | 49 (92.5%) |
| **Age Range** | |
| 20-29 | 10 (18.9%) |
| 30-39 | 13 (24.5%) |
| 40-49 | 8 (15.1%) |
| 50-59 | 7 (13.2%) |
| 60-69 | 10 (18.9%) |
| 70-79 | 4 (7.6%) |
| 80-89 | 1 (1.9%) |
