## Supplementary figures and images for "Respiratory virus detection and sequencing from negative SARS-CoV-2 rapid antigen tests"

### Supplementary Figure 1

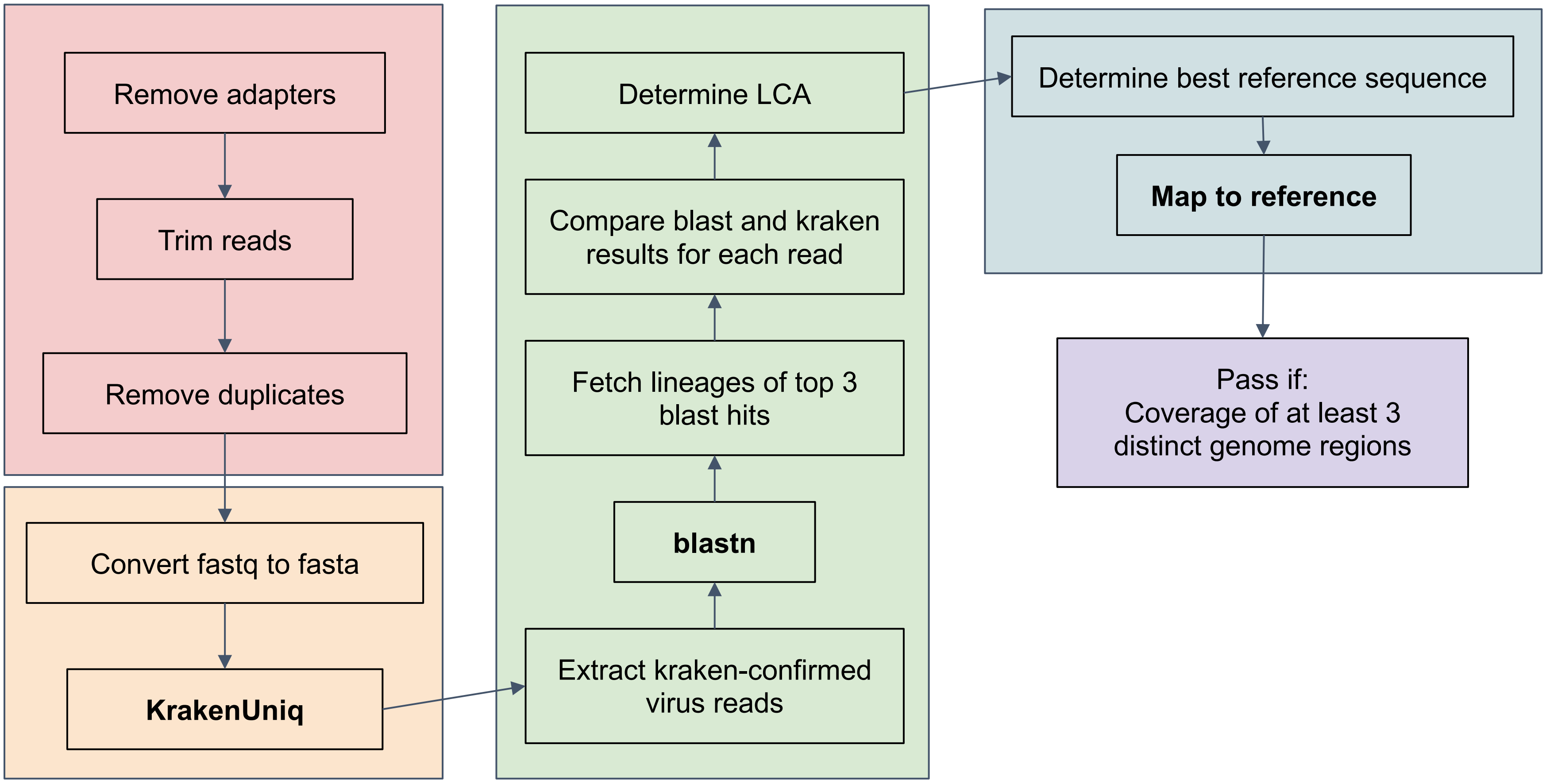

### Supplementary Figure 2

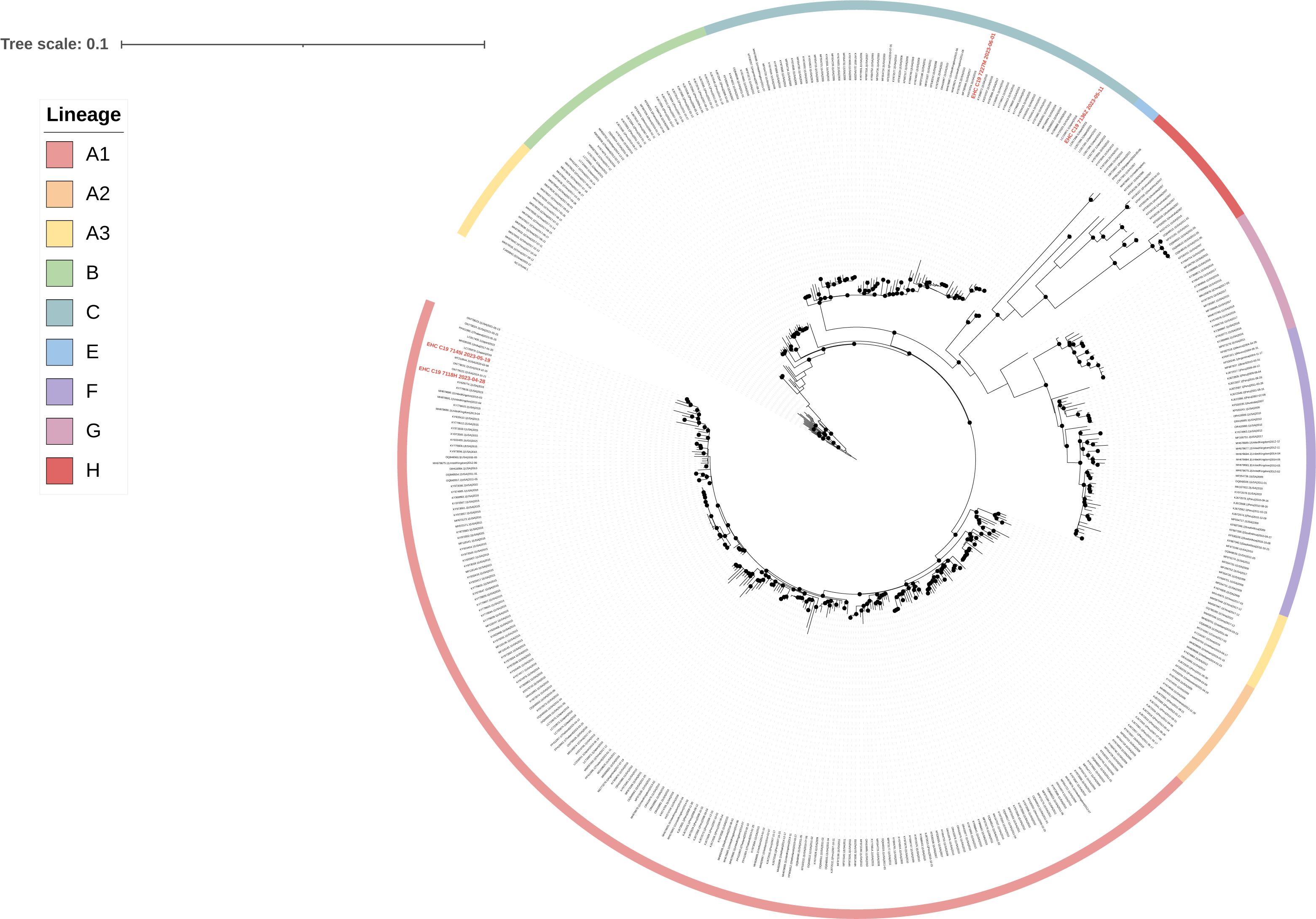
